# “It’s allowed us access to the outside world”: A Qualitative Study of Lifespan Supported Mobility Device Use in Cerebral Palsy

**DOI:** 10.1101/2022.02.17.22269930

**Authors:** Heather A. Feldner, Deborah Gaebler-Spira, Varun Awasthi, Kristie Bjornson

**Affiliations:** Department of Rehabilitation Medicine, University of Washington, Seattle, WA, USA; Shirley Ryan Ability Lab, Chicago, IL, USA; Seattle Children’s Research Institute, Seattle, WA, USA

## Abstract

**Aim:** The overarching aim of this research was to 1) Understand the mobility experiences, supported mobility device (SMD) use, and desired participation outcomes of people with cerebral palsy (CP) across the lifespan; and 2) Describe how perspectives of rehabilitation care and professional resources may influence mobility decision-making processes and outcomes. The aim of this study was to understand the lived experience of SMD provision and use with a focus group guide co-developed by stakeholders.

**Methods:** Focus groups were conducted with 164 participants (people with CP, caregivers, and healthcare providers) across four US cities. Sessions were audio-recorded, transcribed, and analyzed using constant comparison.

**Results:** Six themes emerged. Five presented across all stakeholder groups: 1) *The System is Broken;* 2) *Equipment is Simultaneously Liberating and Restricting;* 3) *Adaptation Across the Lifespan;* 4) *Designed for Transport, not for Living;* and 5) *Sharing Our Stories and Sharing Resources*. One theme was specific to healthcare provider groups: Caught in the Middle.

**Interpretation:** This qualitative study underscores the simultaneous value and frustration associated with SMD, and the need to improve connections and resource networks within the CP community to improve SMD design and provision processes across device types and across the lifespan for people with CP.

**What this paper adds:**

- One of the largest qualitative data sets specific to supportive mobility devices across ages and functional levels.
- Results indicate SMD is most often equated with freedom, participation, and independence.
- Frustrations with SMD across the lifespan persist related to design, function, cost, and maintenance.
- Stakeholders in the CP community are seeking greater networking and resource-sharing to enhance SMD provision processes.
- Access to appropriate SMD across the lifespan and the need for systems improvement is critical.

---

Across the world, cerebral palsy (CP) is the most common perinatal motor disability.^1^ Nearly 10,000 children are diagnosed with CP each year and approximately 764,000 people are living with CP in the US.^2^ People with CP experience heterogeneity of motor function, communication ability, cognition, and participation. However, delays in walking and other mobility skills are common.^3^ To facilitate participation, people with CP across the lifespan benefit from supportive mobility devices (SMD) like orthotics, walkers, crutches, or wheelchairs.^3,4^ Such devices are considered essential environmental factors from the holistic lens of the World Health Organization’s International Classification of Functioning, Disability, and Health framework.^3^ Within this framework, current standards of rehabilitation practice include SMD provision as a part of individualized, person/family-centered intervention.^4^ However, provision processes vary across regional and clinical contexts and a lack of resources exist to guide introduction and evolution of SMD throughout the lifespan, especially as ambulatory ability changes over time.^5,6^ In some cases, clinical trends result in delayed provision of wheeled SMD until efforts to promote independent walking are exhausted, despite evidence supporting its benefits.^7,8^

There is a limited understanding of SMD provision and use from stakeholders with lived experience of CP. For example, younger children view their devices as both functional and social, often incorporating them into play schemes.^9,10^ They often consider SMD as an extension of their bodies, which contributes to development of self-concept and identity either positively or negatively depending on contextual messaging about disability and technology.^11,12^ Adolescents with CP embrace multiple modes of mobility based on their activities and the relative in/accessibility of their environments,^13^ looking upon SMD as an opportunity rather than a failure.^9,14^ Adults with CP recognize SMD as a positive facilitator of participation, while simultaneously critiquing elements of design, choice, and financial/environmental accessibility.^15,16^ Caregivers of children with CP report SMD helps reduce their physical and emotional stress and improves their child’s participation, agency, and sleep patterns.^17,18^ Challenges with SMD maintenance and repair, cost, and bulk/size are also frequently reported.^19-21^

Provision and use of SMD is also an important consideration within an overarching context of shared decision-making and person/family-centered care.^5,22,23^ However, it remains largely unknown how SMD decision-making evolves, especially during transition to adulthood.^22,24^ This gap is concerning, considering that estimated care costs across the lifespan of an individual with CP approximates 1 million dollars,^25^ including SMD. Based on cost projections of SMD alone for people with CP up to age 21, ambulatory individuals (GMFCS level I to III) can have lifetime costs up to 68,000 (+/-20%), with costs for individuals at GMFCS level IV increasing to $90,000 (+/-20%), largely due to SMD needs.^26^ Additionally, high rates of device abandonment lead to needless SMD expenditures and cost increases.^27^ Fiscal implications from the perspective of stakeholders with lived experiences of CP, however, have not been widely studied.

These factors were highlighted by nearly 50 stakeholders during Research CP, a recent participatory action initiative which included webinars, consensus building, and an in-person workshop to address priorities for person-centered CP research.^28^ A better understanding of SMD impact is critical to support these priorities, specifically in identifying interventions (including equipment) to maximize functional outcomes and minimize pain and fatigue throughout aging and across GMFCS levels.^28^

A multi-phase, mixed-methods study was conducted in which the overarching objectives were to 1) Understand the mobility experiences, SMD use, and desired participation outcomes of individuals with CP across the lifespan; and 2) Describe how healthcare provider perspectives and professional resources may influence mobility decision-making processes and outcomes in people with CP and their caregivers. The first phase of the study consisted of a Delphi consensus-building process with nine stakeholders from the CP community to co-develop and prioritize questions and topic areas for an SMD-focused qualitative protocol. This paper describes the second phase of our study, in which the deployment and analysis of this protocol took place.

## Methods

This phenomenological study was conducted with institutional approval from [institution removed for review] Institutional Review Board (#1490). Prior to participating, participants provided written consent and/or permission for all research procedures. All names used are pseudonyms.

### Participants and Setting

Individuals and family dyads (individual with CP + caregiver) and healthcare providers were recruited using purposive sampling across four US cities with regional CP care centers: Boston, Chicago, Los Angeles, and Seattle. Site ‘champions’ were identified through professional networks to assist in procuring space for study procedures and aid in local recruitment (posting of study flyers and email distribution to patient registries or listservs). Potential participants directly contacted a study coordinator, who conducted eligibility screening, study enrollment and scheduling, and follow up. Inclusion and exclusion criteria can be found in Table 1.

**Table 1.** Study Inclusion and Exclusion Criteria

| Criteria | Description |
| --- | --- |
| Inclusion | 1. Individuals with CP of any age and with functional skills consistent with GMFCS levels II through V<br>2. Individuals 18 yrs of age and older who identify as family members or caregivers of individuals with CP<br>3. Licensed or certified healthcare providers (therapists, physicians, assistive technology professionals, etc.)<br>*For individuals under the age of 18, or for individuals over the age of 18 with limited ability to consent due to cognitive status, a parent or legal guardian was required to provide consent for themselves and permission for their child to participate in the study. Verbal assent was sought and recorded for all participants over the age of seven years, even in cases where cognitive status required caregiver permission for study participation. |
| Exclusion | 1. Participant has not had any prior experience with SMD |

### Study Team and Positionality

The research team consisted of two PhD trained pediatric physical therapists, each with between 20-30 years of clinical and research experience with individuals with CP and their families, a physiatrist with over 40 years of clinical and advocacy expertise within the CP community, and an experienced study coordinator. While all members of the research team have worked extensively with the CP community professionally, none identify as having lived experience with CP. For this reason, it was critical to convene a nine-member Stakeholder Advisory Panel, which consisted of individuals with ambulatory and non-ambulatory CP, caregivers, and healthcare providers to co-develop and guide the study as well as participate in data analysis and interpretation activities.

### Study Procedures

Focus groups are a valuable way to gain perspectives from people with homogenous experiences, encouraging participants to elucidate their views and express dis/agreement in a group dynamic.^29^ Focus groups consisting of 6-8 individuals or family dyads were carried out by two research team members with expertise in qualitative methods. Attempts were made to stratify focus groups by age and GMFCS levels (age bands of 0-7 yrs, 8-18 yrs, and 21+ years; GMFCS II-III, GMFCS IV-V), so participants were more likely to have some crossover in SMD experience. Based on participant demographics this was not always possible for GMFCS level, age stratification was largely successful. Professional focus groups took place separately from participants with CP, without stratification needed.

Focus groups were held in accessible community locations at each study site. Researchers established ground rules (i.e. validation of all perspectives and experiences, sharing without interruption, silencing cell phones, maintaining confidentiality, plus additional rules agreed upon by each group), and facilitated discussion. Researchers used the semi-structured focus group guide co-developed by the Stakeholder Advisory Panel during the first phase of the study using a Delphi consensus technique.^30^ An excerpt of the guide is included in Figure 1. Focus group sessions lasted between 60-90 minutes and all sessions were audio recorded. Participants were issued a $30 gift card as compensation for their time and expertise.

**Figure 1.**
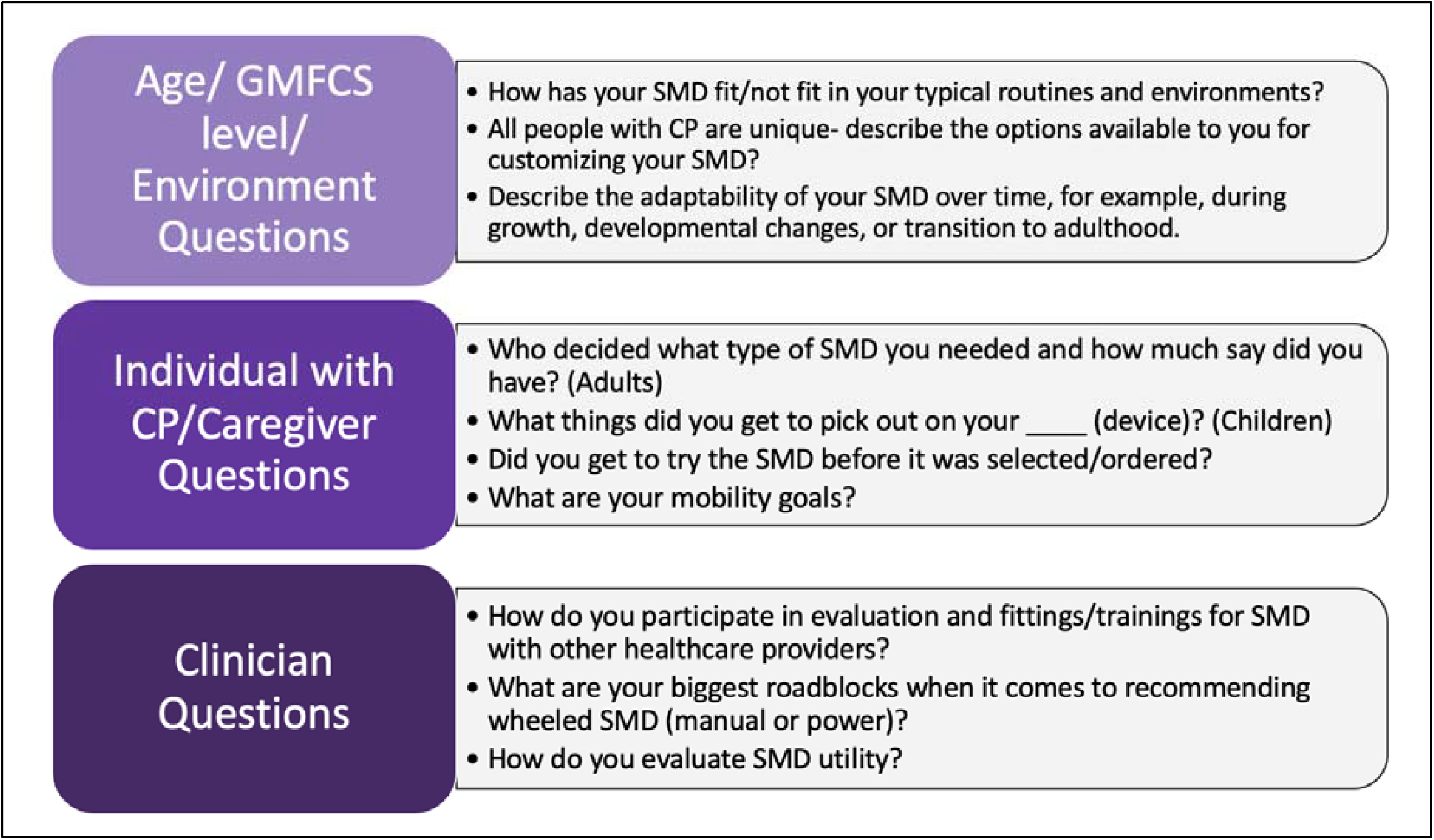
Sample of Semi-Structured Focus Group Questions. This figure shows a sample of semi-structured focus group questions asked at each session. These questions were written and prioritized by a 9-member Stakeholder Advisory Panel representing the CP community during a previous study activity and are divided based on the openended categories used to generate the questions.

### Data Analysis

Focus group recordings were transcribed verbatim and coded. Independent, hand-coding of transcripts was conducted by the research team and the Stakeholder Advisory Panel. A constant comparative method was employed to create open codes, narrow to focused codes, and ultimately determine data saturation to derive central themes.^29^ Following initial independent coding rounds, the three lead researchers met to discuss and refine code groupings until themes emerged, using discussion to resolve disagreement and respond to questions until 100% agreement was reached. To ensure rigor and minimize researcher bias, an audit trail was created for transparency, an example coding scheme is included in Figure 2. Additionally, thick descriptions of participant experiences were extracted from the data to ensure context was maintained. Researchers engaged in self-reflection to identify potential biases, and member checking was conducted with all participants to ensure accuracy of the themes and avoid misinterpretation of the data.^29^

**Figure 2.**
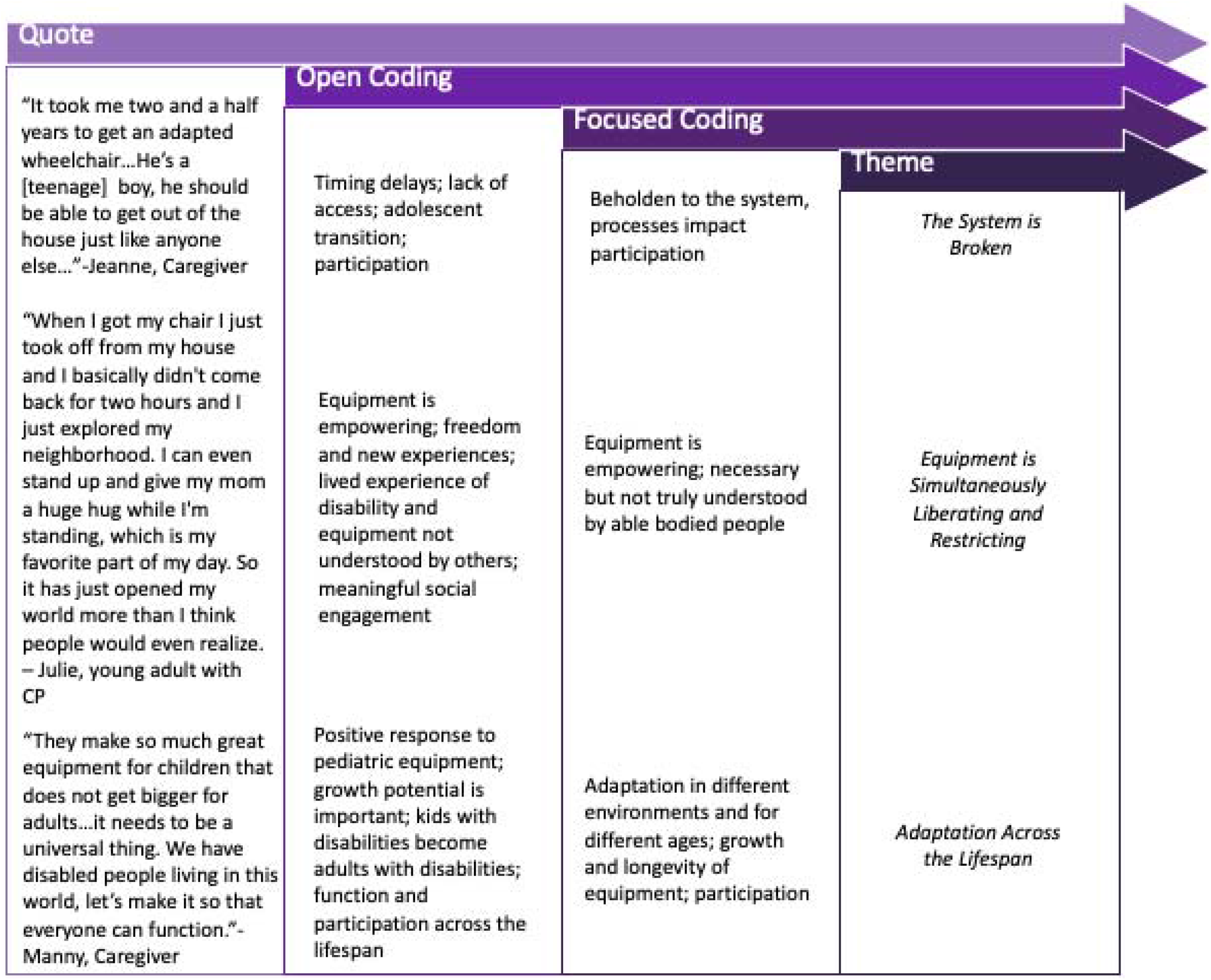
Coding Process and Evolution of Thematic Analysis. *Sample Coding Scheme* This figure depicts the coding scheme, which is part of the qualitative audit trail to ensure rigor in qualitative data analysis and interpretation. The quotes were first coded into open codes by multiple researchers, then condensed into focused codes, with discussion to resolve any disagreement as themes emerged from the focused codes.

## Results

A total of 164 participants took part in 24 focus groups. This included 68 individuals with CP (ages 3-68), 74 caregivers, and 22 healthcare providers (physicians, occupational and physical therapists, and Assistive Technology Professionals). See Table 2 for participant demographics.

**Table 2.** Participant Demographics

|  | Seattle | Chicago | Los Angeles | Boston | All site Total |
| --- | --- | --- | --- | --- | --- |
| **Participants** |  |  |  |  |  |
| Caregivers (consented) | 16 | 23 | 15 | 20 | 74 |
| Professionals (consented) | 10 | 8 | 2 | 2 | 22 |
| Participants with CP (consented) | 0 | 7 | 8 | 0 | 15 |
| Participants with CP (via Caregiver Permission) | 16 | 15 | 7 | 15 | 53 |
| Total | 42 | 53 | 32 | 37 | 164 |
| **Age Range, Participants with CP (either consented or via Caregiver permission)** |  |  |  |  |  |
| 0--6 | 7 | 0 | 0 | 3 | 10 |
| 7--12 | 5 | 5 | 0 | 8 | 18 |
| 13--20 | 4 | 9 | 3 | 4 | 20 |
| 21+ | 0 | 9 | 11 | 0 | 20 |
| Total | 16 | 23 | 14 | 15 | 68 |
| **Racial and Ethnic Identity (all participants)** |  |  |  |  |  |
| Caucasian | 33 | 48 | 28 | 36 | 145 |
| Black or African American | 7 | 3 | 1 | 1 | 12 |
| Asian | 0 | 0 | 3 | 0 | 3 |
| Alaska Native/Pacific Islander | 2 | 0 | 0 | 0 | 2 |
| Middle Eastern | 0 | 2 | 0 | 0 | 2 |
| Hispanic/Latino | 2 | 6 | 2 | 0 | 10 |
| Non-Hispanic/Latino | 40 | 47 | 30 | 37 | 154 |

Six themes emerged from the data. Five were present across all stakeholder groups: 1) *The System is Broken;* 2) *Equipment is Simultaneously Liberating and Restricting;* 3) *Adaptation Across the Lifespan;* 4) *Designed for Transport, not for Living;* and 5) *Sharing Our Stories and Sharing Resources*. One additional theme emerged specific to the healthcare provider groups: *Caught in the Middle*. These themes are discussed below, with corresponding participant quotes found in Table 3.

**Table 3.** Participant Quotes

| Quote Number | Representative Quote | Theme |
| --- | --- | --- |
| 1 | *"One of the **biggest issues is other people**. Outside sources, insurance companies, these people sitting in offices **telling my daughter what she can or can't have or what she will and won't be able to use**. Whereas PTs who work their butts off to get these letters written, and **my parental input saying what we will and won't use at home** and what will be successful, just for somebody in an office to say, "no you can't, because we already gave you one thing.""*- Daphne*, Caregiver | The System is Broken |
| 2 | *"It took me **two and a half years** to get an adapted wheelchair...He's a [teenage] boy, **he should be able to get out of the house just like anyone else...**"*-Jeanne, Caregiver | The System is Broken |
| 3 | *"If [our equipment] breaks, it's like a whole month to come and repair that part, and **my son has to miss his school**."* – Stephanie*, Caregiver | The System is Broken |
| 4 | *"I'm just saying how ridiculously expensive everything is. **A wheelchair costs as much as a car**. Come on, it's ridiculous!"*-Tasha, Caregiver | The System is Broken |
| 5 | *Why can't we get a bike approved? A bike is a necessity for normal growth and learning and interaction with other peers. All these kids are sitting on the sideline and they shouldn't be. We're holding them back educational and physically and socially and they're part of society and have a lot to give back. We have to get better and a bike doesn't have to be \$5,000."* – Carlos*, Caregiver | The System is Broken |
| 6 | *"And some things, **I don't know if I would have chosen...**I don't know if we would have chosen that stander over another one, but we couldn't try them out, and it is hard when you spend crazy amounts of money and then you have it and you can't return it. I wish that there was a place like that where you could just go try out all this stuff and be like, "Oh, that would work perfect.""* - Alyssa*, Caregiver | The System is Broken |
| 7 | *"When I got my chair I just took off from my house and I basically didn't come back for two hours and **I just explored my neighborhood**. I can even stand up and **give my mom a huge hug** while I'm standing, which is my favorite part of my day. **So it has just opened my world more than I think people would even realize**."* – Julie*, young adult with CP | Equipment is Simultaneously Liberating and Restricting |
| 8 | *"**Sometimes he forgets he has a disability**, kind of, because In class they give them some task, like be the line leader and stuff, and he gets involved and when they move from the classroom to the gym room which is a distance, **he moves around with the crowd**, gets there [with his SMD]. It's allowed us access to the outside world"* -Cheryl*, Caregiver | Equipment is Simultaneously Liberating and Restricting |
| 9 | *"The wheelchair works in sand dunes and my walker works really well, a bit better on the grass and* | Equipment is Simultaneously |
|  | *in the water too!" -Chris\*, child with CP* | *Liberating and Restricting* |
| 10 | *"They decided to put braces on me...we rode to school in taxis, so **in the taxi home I would start unbuckling the braces...** I would jump out, drop the braces on the front lawn, and go in the house. **As a little kid, I was setting myself free.**" – Sabrina, adult with CP* | *Equipment is Simultaneously Liberating and Restricting* |
| 11 | *"[One of the challenges] with the crutches and the walker for me...I won't be able to have my hands free. Because you can't really do anything unless you stop, put them down, hold on to something...then you can function" -Monique\*, young adult with CP* | *Equipment is Simultaneously Liberating and Restricting* |
| 12 | *"I need a chair that's going to be able to **adapt to my needs in that environment**. For example, sometimes I cannot fit under the desk in the classroom" –Alexis, young adult with CP* | *Adaptation Across the Lifespan* |
| 13 | *"They make so much **great equipment for children that does not get bigger for adults...**it needs to be a universal thing. We have disabled people living in this world, **let's make it so that everyone can function.**"-Manny\*, Caregiver* | *Adaptation Across the Lifespan* |
| 14 | *"Equipment never keeps up with the pace of the... either the development of the child or the growth of the child, which are two different aspects." -Charlotte, OT and Assistive Technology Professional* | *Adaptation Across the Lifespan* |
| 15 | *"You can't manufacture a product for everyone because every single person in this room has a different need or set of needs. They need to listen and they need to observe and they need to measure and they need to really create the implement for the need of that individual." – Jack\*, Assistive Technology Professional* | *Adaptation Across the Lifespan* |
| 16 | *"I would want my equipment to **express my personality** and things that I love" – Monique\*, Young Adult with CP* | *Designed for Transport, not for Living* |
| 17 | *"They limit our colors or they **limit our attractiveness** – Cindy\*, adult with CP* | *Designed for Transport, not for Living* |
| 18 | *"I don't think that the companies think about the whole picture. **They look at the immediate effects.** That chair gets you from point A to point B on a flat surface indoors. That's it. **They don't look at the big picture of your daily life.**"- Jocelyn\*, Caregiver* | *Designed for Transport, not for Living* |
| 19 | *"**Flying** would be kinda cool!" – Malik\*, child with CP* | *Designed for Transport, not for Living* |
| 20 | *"My wife and I both had **careers enhanced, lives changed** by being the parents of a child with CP. She founded a non-profit to teach all the things she has learned..." - Harvey\*, Caregiver* | *Sharing our Stories and Sharing Resources* |
| 21 | *"We're going to the beach. It was a state park, and on their website they said, "Beach wheelchairs available upon request. Just talk to the ranger." We pulled in, talk to the ranger, they said, "no* | *Sharing our Stories and Sharing Resources* |
|  | *problem." They brought it out. **It was the best thing ever, because he could go for a walk with us or just get to the spot that we were going to sit on the beach. It was fabulous.**" – Julia\*, Caregiver* |  |
| 22 | ***"We're getting a lot of information from each other. Through word of mouth. And it should be coming from the professionals. Yeah, you can advocate all you want, but it shouldn't take two people getting in a room together and talking [about equipment] for a person to say, "Okay, I need this to better my life or better my situation."** – Alexis\*, young adult with CP* | *Sharing our Stories and Sharing Resources* |
| 23 | ***"The hospitals or clinics are not willing to pay for tech support to have things built and adjusted and repaired... some of the manufacturers are in such a crunch that they can't provide very many trial items, or only in one size. So I feel like there's been a strangle hold on some of us"** – Cynthia\*, OT* | *Caught in the Middle* |
| 24 | ***"It's really difficult to manage expectations, in this field we're dealing with a lot of very custom approaches or outcomes and not being able to physically test a lot of those custom interventions, but still speak to those, and then get insurance to agree that that is going to be the right way."** – Keith\*, Assistive Technology Professional* | *Caught in the Middle* |

\*All names are pseudonyms

### The System is Broken

This theme described challenges faced by people with CP and caregivers as they navigate SMD procurement, use, and maintenance. Participants recognized they must work within a flawed system often regulated by unique policies based on state of residence, types of funding available, and the knowledge, preferences, and availability of individual providers. Many participants expressed frustration with a consistent cycle of SMD denials and appeals despite advocacy by their rehab teams (Table 3, Quote 1). Lengthy delays between evaluation and delivery or for repairs to essential SMD were frequently reported (Table 3, Quotes 2 and 3). Participants highlighted issues of cost, citing a system which labels SMD as ‘specialized’, resulting in significant price inflation that impacts participation (Table 3, Quotes 4 and 5). A lack of knowledge about different SMD options and a lack of trial equipment were also common (Table 3, Quote 6).

### Equipment is Simultaneously Liberating and Restricting

The second theme described perspectives of SMD as critical facilitators of independence, agency, and self-concept throughout the lifespan, while simultaneously noting SMD-related barriers to participation. Nearly all participants shared their excitement about the functional and social freedom and inclusion facilitated by SMD (Table 3, Quotes 7 and 8). Another common finding was the recognition that multiple forms of SMD were essential to navigate different environments and situations (Table 3, Quote 9). Despite these clear benefits, participants simultaneously noted restrictive aspects of their SMD, including limited access to certain activities or environments (Table 3, Quotes 10 and 11).

### Adaptation Across the Lifespan

The third theme described the need for both SMD and environments to better adapt and respond to individual needs across the lifespan. For example, participants described practical challenges such as fitting under desks or in workspaces (Table 3, Quote 12). Participants also noted frustration with the lack of SMD carrying over during the transition to adulthood (Table 3, Quote 13). As one participant stated, “I’ve had CP all my life, it’s not going away just because I’m an adult now.” (Mia, adult with CP). Other participants discussed challenges related to SMD keeping pace with growth and/or development (Table 3, Quote 14). Participants explored tensions between adaptability and the need for multiple types of SMD, agreeing that while adaptation is critical, there will always be a simultaneous need for custom SMD to meet the unique needs of individuals with CP (Table 3, Quote 15).

### Designed for Transport, not for Living

The fourth theme described frustration with perceived lack of design and aesthetic innovation for SMD as an essential part of life. It also encompassed creativity and innovative ideas about SMD. For example, participants discussed their desire for SMD to reflect their personality (Table 3, Quote 16). They noted that there is limited choice when it comes to aesthetics, which was at times incongruent with their desire to express themselves with their SMD as an extension of their body (Table 3, Quote 17). Participants also agreed that insurance companies and other outsiders viewed most SMD as a means of transport, rather than a key means of participation in family and community activities (Table 3, Quote 18). Young children expressed their biggest wishes for their SMD design and function, including flying, temperature control, or a self-cleaning wheelchair (Table 2, Quote 19).

### Sharing Our Stories and Sharing Resources

The fifth theme represented a call for voices to be heard more explicitly, to receive and provide support for others, and create a means of centralized information sharing to empower the community. For example, some caregivers noted that advocacy efforts that began as a parent of a child with CP turned into career opportunities (Table 3, Quote 20). Other participants shared their SMD successes to help others with identified barriers, such as seamless access to a beach chair for a day trip or grant funding opportunities for a needed SMD item (Table 3, Quote 21). Most participants noted some degree of isolation in navigating the complexities of life with CP. They recognized a lack of community and shared knowledge, especially early on in their CP journeys. Simple activities like the SMD focus groups were a powerful way to build stronger communities and learn from each other (Table 3, Quote 22).

### Caught in the Middle

The final theme was specific to healthcare providers. This theme described being caught in between wanting to provide optimal care and SMD to their clients, yet recognizing barriers such as SMD design and access, constraints of funding limitations, increasing regulatory climates, and reduction of specialty seating clinics across the US. For example, professionals recognized the challenges of preserving salary support for ATPs, appropriate trial items, and having to contend with frequent appeals and delays in ordering SMD (Table 3, Quote 23). They also described the fine balance between empowering families, working within funding constraints, and relying on clinical experience for customized SMD solutions that may not yet have a significant evidence-base to draw from (Table 3, Quote 24). Managing expectations within this complex professional climate was noted as one of the field’s most significant challenges.

## Discussion

This study, the second phase in a mixed-methods, multi-phase research project, conducted focus groups across a large sample of people with CP, caregivers, and healthcare providers to understand experiences of SMD provision and use across the lifespan. Resulting themes add rich context, highlighting the complex landscape of SMD previously reported for people with CP and their caregivers.^11,13,15^ Findings offer new insights regarding lived experience and healthcare provider perceptions that may inform collaborative care, but have rarely been addressed in the literature to date.^4,5^

In particular, Themes 1 and 2 align with findings reported and research priorities established during Research CP, indicating that current standards of practice may not meet everyday needs of people with CP across the lifespan aiming to maximize participation and minimize pain and fatigue.^28^ Theme 2 results also reflect existing literature describing how young children and adolescents perceive SMD as positive extensions of their bodies that support social-emotional development, along with decreased caregiver stress.^9,12,13,18^ This theme was mixed, however, with responses also mirroring evidence documenting consistent challenges with repair and maintenance, barriers to participation, and device design.^7,15,16,21^ Themes 3 and 4 correspond with the identified need for ongoing SMD development across the lifespan and reported frustration with a lack of adaptable SMD across environments and stages, particularly during transition to adulthood.^3,28^

Participant responses in Theme 5 provide novel evidence to enhance person/family-centered care through sharing stories and resources about SMD experiences.^5^ This spontaneous networking was a powerful and unexpected occurrence across multiple focus groups, pointing to the need for greater CP community engagement in general, but particularly to fill a knowledge gap related to SMD. Similarly, across Themes 1 and 6, participant responses described frustrations with policy factors that influence access and customizability for SMD users, another knowledge gap that has not been widely explored in CP research to date.^26^

From an overarching perspective, study results point to focus areas across research, clinical practice, and policy/advocacy where SMD provision and use experiences of people with CP may be enhanced. It also points to the need for additional participatory and cost effectiveness research that will shift the design and provision of SMD from transport to living (Theme 4) and may address barriers and limitations noted by participants. Doing so successfully will require ‘champions’ across all stakeholder groups locally and nationally as well as leveraging knowledge of how shared-decision making can enhance the cost effectiveness and satisfaction of SMD support across the lifespan.^24,27^

### Limitations and Future Directions

This study has several limitations. First, while a national sample of participants was recruited, selection bias may have skewed responses, since participants were mostly white and self-selected to take part in SMD focus groups. Second, because policy and funding implications differ widely state to state, and there is a lack of understanding about how funding agencies make SMD decisions, the absence of funding and policy representatives in our study is a clear limitation that will be rectified in future work. Third, the presence of a researcher with professional SMD experience has the potential to produce acquiescence bias, though the team attempted to mitigate this through unconditional positive regard as well as seeking out multiple and discordant/outlying viewpoints.^29^ Finally, the large sample size resulted in a substantial qualitative data set. Though Stakeholder Advisory Panelists were provided explicit instructions for assisting with data coding and all codes were reviewed by the primary research team, coding idiosyncrasies among a large analysis team could have impacted thematic results. This limitation was mitigated by creating an audit trail to document all coding decisions as well as sharing thematic results and descriptions with participants during member-checking.^29^

Shorter term future work includes conducting focus groups with funding agency and policy representatives. Long-term work will include the implementation of a national ‘smart survey’ stratified by age and GMFCS level to inform the development of a clinical algorithm that supports SMD provision and educates stakeholders about common barriers and potential solutions to optimize timing and provision of SMD across the lifespan of people with CP.

## Conclusion

This study represents one of the largest SMD-focused qualitative studies to date within the CP community. Results demonstrate that qualitative Inquiry is a powerful way to foreground the lived experience of CP to improve understanding of SMD-specific experiences and needs. Participants were eager to take part in focus groups and have their voices heard, with impromptu community building and resource-sharing occurring as an unexpected outcome. This study indicates that the timing and provision of SMD should be a dynamic, interactive, and shared decision process between the individual and/or family, and healthcare providers, involving a systematic, process-oriented approach generated directly from the experiences and needs of the CP community.

## Supporting information

SRQR Checklist

## Data Availability

All data produced in the study are available upon reasonable request to the authors.

## List of Abbreviations

SMD: Supportive Mobility Device
CP: Cerebral Palsy

## Acknowledgements

The authors would like to thank the Stakeholder Advisory Panel, all of whom were critical in carrying out this work: Susan Johnson-Taylor, Tim Caruso, Dave Pruitt, Holly Wakefield, Karla Lynch, Jan Brunnstrom-Hernandez, Lauren Rosen, Sariya Rashid, and Candi Styer. We would also like to thank the focus group site champions who assisted with recruitment, focus group facilities, and accommodation recommendations: Jessica Pedersen, Josephine Boggs, Patricia Herbst, Ben Shore, Jodie Shea, Eileen Fowler, and Marcia Greenberg. Finally, we would like to thank all our focus group participants, young and old. Your lived experiences are the heartbeat of this work and will continue to inform our future research, teaching, and advocacy.

